# Acute intermittent hypoxia enhances volitional elbow strength, and modulates spatial distribution of muscle activation patterns in persons with chronic incomplete spinal cord injury

**DOI:** 10.1101/2022.08.23.22278497

**Authors:** Babak Afsharipour, Gregory EP Pearcey, W Zev Rymer, Milap S Sandhu

## Abstract

**Background:** Acute intermittent hypoxia (AIH) is an emerging technique for facilitating neural plasticity in individuals with major neurological deficits. In people with chronic incomplete spinal cord injury (iSCI), a single sequence of AIH enhances motor function such as hand grip strength and ankle plantarflexion torque, but the underlying mechanisms are not yet clear.

**Objective:** To examine how AIH-induced changes in magnitude and spatial distribution of electromyography (EMG) activity over the surface of the biceps and triceps brachii muscles contributes to improved strength.

**Methods:** Seven individuals with iSCI visited the laboratory on two occasions, at least a week apart, and received either AIH or Sham AIH intervention in a randomized order. AIH consisted of 15 brief (∼60s) periods of low oxygen (fraction of inspired O_2_ = 0.09) alternating with 60s of normoxia, whereas Sham AIH consisted of repeated exposures to normoxic gas mixtures. Muscle activity of biceps and triceps brachii was recorded with high-density surface EMG during maximal elbow flexion and extension contractions. We used these EMG recordings to generate spatial maps which distinguished active muscle regions prior to and 60 minutes after AIH or Sham AIH.

**Results:** After an AIH sequence, elbow flexion and extension forces increased by 91.7 ± 33.5% and 51.7 ± 21.9% from baseline, respectively, whereas there was no difference after Sham AIH exposure. Changes in strength were associated with an altered spatial distribution of EMG activity and increased root mean squared EMG amplitude in both biceps and triceps brachii muscles.

**Conclusions:** These data suggest that altered motor unit activation profiles may underlie improved volitional strength after a single dose of AIH and warrant further investigation using single motor unit analysis techniques to further elucidate mechanisms of AIH-induced plasticity.

## Introduction

Most spinal cord injuries (SCI) are incomplete, but the majority result in long-lasting motor impairments. Despite improvements in therapeutic modalities over time, no single therapy has proven to be overly effective for overcoming these impairments. The combination of task-specific training modalities coupled with neuromodulation approaches is most likely to have the greatest impact on functional recovery ^1,2^. Mild doses of acute intermittent hypoxia (AIH), which consist of repeated but brief periods of breathing low oxygen gas mixtures, has gained attention in recent decades due to its facilitatory effects on respiratory and non-respiratory motor function ^2,3^. AIH triggers the release of serotonin, and increases the synthesis of brain-derived neurotrophic factor (BDNF), one of the most important regulators of neuroplasticity ^3–5^. Higher ‘doses’ of hypoxia are known to contribute to pathologies in the central nervous system (i.e., neuro-inflammation) ^6^, but also provide additional evidence for neuroplasticity. Lower ‘doses’ (i.e., AIH), on the other hand, seem to circumvent the pathologies related to more severe hypoxia, and promote motor plasticity and therapeutic benefits ^7^.

Our lab has shown that a single sequence of AIH (i.e., 30 minutes) can significantly increase function in the somatic motor system. This is evidenced by an increase in strength of muscles acting at the ankle joint ^8–10^ as well as wrist/hand function ^11^. Despite the known mechanisms responsible for AIH-induced neuroplasticity in the neural control of respiratory muscles observed in the laboratory of Mitchell and colleagues ^4,5,12–17^. However, there is less known about the mechanisms responsible for AIH-induced neuroplasticity in the somatic motor system, especially in humans. Indeed, corticospinal synaptic plasticity is potentially enhanced after AIH ^18^, but many other sites are probable contributors as well.

Along with strength changes, there have been observed changes in the amplitude and coordination of single differential electromyography (EMG) recorded over the agonist and antagonist muscles of the ankle following AIH ^8,10^, suggesting that activation and/or excitability of motoneurons may also be enhanced by AIH. There are, however, many concerns with the analysis of single differential EMG, because the small area of recording has the potential for error of both types I and II ^19^ that can potentially be avoided with the use of high-density surface EMG (HDsEMG) recorded over a larger surface of the muscle of interest.

In this study, we examined how AIH affects strength and muscle activity in proximal arm muscles in people with chronic incomplete SCI. More specifically, we recorded HDsEMG from the elbow flexors (biceps brachii) and extensors (triceps brachii) during maximal voluntary elbow flexion and extension torque generation, respectively. We hypothesized that, since AIH is effective in increasing volitional strength in muscles acting at the ankles and wrists of spinal cord injured patients, we should observe increases in strength about the elbow too. We also hypothesized that we would observe increased intensity and the area of muscle activation after AIH. Use of high-density surface EMG recordings will also allow us to determine whether increased muscle forces are attributable to an overall increase in excitation of the motor pool (in which case the shape of the EMG map should stay approximately constant), or whether other sections of the motoneuron pool are engaged, perhaps because these sections are affected by the spinal cord lesion and/or contain different motor unit types.

## Methods

### Participants

Seven individuals (48.1 ± 11.79 years; 5 males, 2 females) with incomplete spinal cord injuries in the cervical region ranging from C4-C7 participated in the study (see table 1 for characteristics of study participants). All participants provided written informed consent to the experimental procedures, which were approved by the local ethics committee at Northwestern University and performed in accordance with the Declaration of Helsinki (IRB protocol STU00201602).

**Table 1.** Participant characteristics.

| **Subject** | **Sex** | **AIS** | **Level** | **Age** | **Time Since Injury** |
| --- | --- | --- | --- | --- | --- |
| s01 | M | D | C5 | 61-65 | 3 |
| s02 | M | C | C4-C7 | 46-50 | 11 |
| s03 | M | D | C4-C7 | 46-50 | 4 |
| s04 | F | C | C4-5 | 36-40 | 10 |
| s05 | M | D | C4-C5 | 46-50 | 3 |
| s06 | M | C | C5-C6 | 51-55 | 16 |
| s07 | F | D | C5-C7 | 21-25 | 2 |

### Experimental Protocol

All participants performed two experimental sessions;

1. AIH, which was comprised of signals being recorded before (PRE) and 60 minutes after the AIH intervention (60mPOST), and
2. SHAM AIH, which was identical to the first except that the intervention was a sham intervention. The session type (AIH or SHAM) was randomly chosen and was blinded to the participants at the time of experiments.

For the duration of each experiment, participants were seated comfortably in a Biodex chair, with the elbow in 120° flexion, shoulder in 30° flexion and 35° abduction, and the forearm pronated to 45°. To ensure force isolation, we used orthopedic casting around the forearm, and secured the casted forearm in a custom-built fixture centered at the wrist. The fixture was equipped with a six degree-of-freedom load cell (Delta from ATI, NC, USA) to record torques generated at the wrist (see Fig. 1).

**Figure 1:**
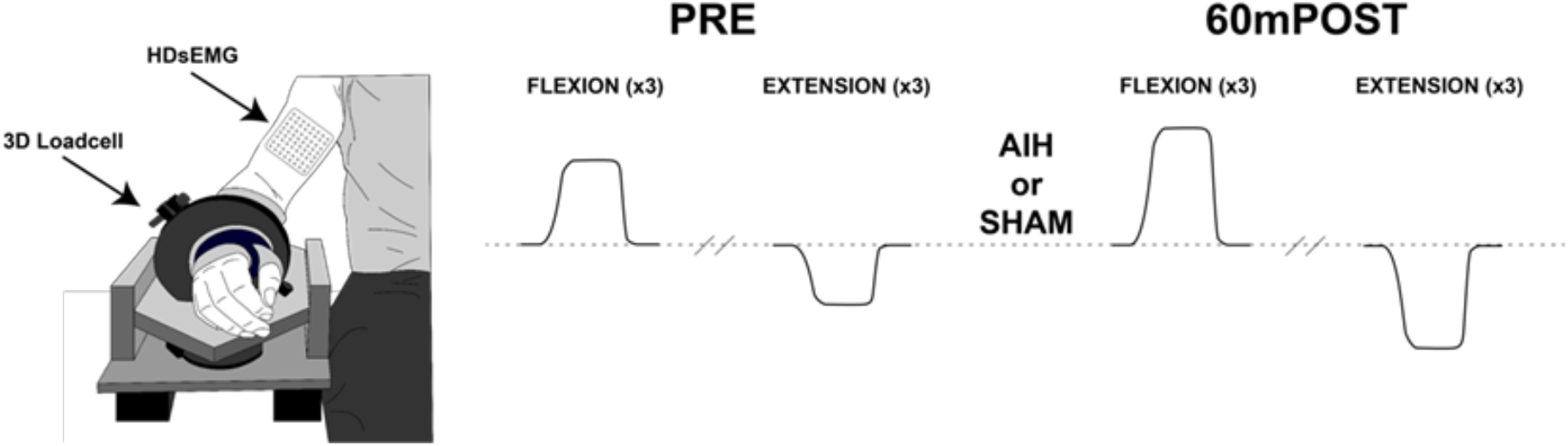
**The experimental setup and timeline used to record forces at the elbow and high-density surface electromyography (HDsEMG). Participants performed three maximal elbow flexion and extension contractions during PRE and 60 minutes POST (60mPOST) during either acute intermittent hypoxia (AIH) or sham AIH (SHAM)**.

We recorded high density surface electromyography (HDsEMG) data using a grid of electrodes applied to biceps brachii (BB; long and short heads), and triceps brachii (TB) muscles of the less affected arms of the participants. The 64-channel HDsEMG channels were arranged in 8 rows and 8 columns (8×8 electrode grid) and electrodes where equally spaced in rows and columns by 8.5 mm (inter electrode distance: IED). One grid (8×8) was used for BB recording and one grid was dedicated to collect signals from TB.

To identify the long and short heads of BB on the grids, we designated the line joining the acromion and the distal Biceps tendon insertion to be the medial-lateral center of the grid. All signals were recorded in monopolar configuration, where the instant amplitude of each electrode was measured relative to subjects’ lateral epicondyle. We measured the relative distance of the grid location to the bony landmarks such as acromion, medial and lateral epicondyles to replace the electrode grid at the same place for consistency among the two signal recording sessions (AIH and SHAM), one week apart.

At each time point in each recording session (AIH or SHAM, PRE or 60mPOST), the HDsEMG channels were recorded during sustained maximal voluntary isometric contractions (MVC) in the elbow flexion, and elbow extension directions. Investigators monitored real-time visual feedback about participant force trajectories (i.e. resultant force direction and magnitude) to ensure the intended actions were performed.

### Acute intermittent hypoxia (AIH) intervention

After PRE measures of force and HDsEMG in both sessions, we secured a latex-free full non-rebreather mask onto the participant using a custom neoprene head strap (Hypoxico INFO). The AIH intervention was administered using a hypoxia generator (Model HYP-123, Hypoxico Inc., New York, NY, USA), which was used to provide 60 s of 9% O_2_ (Fraction of inspired air [FIO_2_]: 0.09) interleaved with 60 s of 21% O_2_ (FIO_2_: 0.21). We repeated the delivery of hypoxia and normoxic air mixtures 15 times per session, for a total of 30 min. Continuous pulse oximetry (Smith Autocorr Plus, Smiths Medical, Dublin, OH, USA) was used to ensure oxygen saturation (SpO_2_) fell to a nadir between 80 and 87% during hypoxic exposures. The SHAM intervention consisted of alternating exposures of normoxic air (i.e. 30 minutes of 21% O_2_).

### Safety monitoring

We monitored several key variables of cardiorespiratory function (GE Dash 4000 Monitor, Chicago, IL, USA) throughout the protocols to ensure that heart rate (50–160 beats per min), systolic blood pressure (85–160 mm Hg), and oxyhemoglobin saturation (>75%) remained within safe limits. We also monitored and asked for self-report of headaches, pain, lightheadedness, dizziness, respiratory distress, spasms, and autonomic dysreflexia, before, during, and after treatment.

### High Density Surface Electromyography (HDsEMG)

Before electrode placement, the grid area and the reference electrode area were lightly abraded and cleaned with alcohol pads to increase signal to noise ratio. The reference electrode was placed over the lateral epicondyle. After securing the arm and placing the EMG grid over the biceps and/or triceps. A 128-channel Refa (TMSi; Oldenzaal, NL) recording system with sampling frequency of 2000Hz was used to collect BB and TB HDsEMG signals. The electrode grids remained in the same position for both the before and after the AIH intervention.

### Data processing

All recorded data (HDsEMG & Force) were converted offline to Matlab for further processing. The HDsEMG data was pre-processed before generating the maps. Pre-processing included mean value removal, band-pass filtering, powerline interference attenuation, bad channel detection and interpolation from all recorded channels. For power line removal, we used a spectral line interpolation technique, where the amplitude spectrum and its phase at power line frequency were interpolated by averaging its neighbor spectra lines (2Hz window) in the frequency domain. Bad channel detection within the grid was based on visual inspection in the time and frequency domains. We replaced the bad channels in the grid with the average of all available good neighbor channels within one inter-electrode distance. The numbers of bad channels were limited, and we found 2 - 4 bad channels out of the 128 channels in a few sessions.

After pre-processing, we obtained the single differential (SD) signals along the fiber direction. The root mean squared (RMS) for each channel was computed (see equation 1 below) as the average RMS over 1s non-overlapping epoch windows along the total length of signals.

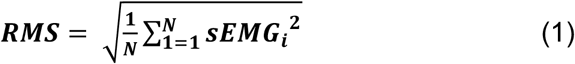

N represents the total samples of a sEMG signal (one channel) and “i” is the sample index. The RMS value of each channel was considered as the amplitude of that channel and was used as intensity of its corresponding pixel in our RMS maps.

The RMS values across each grid were then color coded to represent the map of activity as a heatmap. We segmented the active region of each RMS maps using the watershed segmentation technique to obtain the active region(s) of the RMS maps and extract channels inside the active region(s). We derived the spatial average of the sEMG signals of all active channels for both BB and TB muscles. The spatial averaged sEMG signals were considered as the representative muscle activity during maximal flexion and extension.

Finally, we took a threshold of 75% of the maximal RMS signal from PRE to gain an appreciation of the area of activity that exceeded this threshold in both pre and post. In this case, the number of channels that exceeded this threshold were then divided by the total number of good channels to give a ratio of the grid (i.e. [# of channels > 75% of PRE/total channels]*100% = ratio of active channels as a %).

### Statistical analysis

We used a randomized, double-blinded crossover design to test our hypotheses that a single sequence of AIH enhances maximum elbow flexion and extension strength, and also increases the magnitude and spatial distribution of muscle activity. We assessed differences in each variable independently for each muscle/direction of contraction from baseline and between treatments via Wilcoxon matched pairs signed rank tests. We corrected for multiple comparisons using the Holm-Bonferroni method. Corrected p-values were considered significant at p < 0.05. Mean differences from baseline are reported as means ± standard deviation. In addition, we report Hedge’s G effect size to provide a standardized magnitude of the difference between PRE and 60mPOST after AIH or SHAM intervention.

Next, we determined the relative change in strength and EMG from PRE to 60mPOST AIH for each participant and attempted to identify relationships between changes in EMG and changes in strength using a generalized linear mixed model (i.e., GLMM). This was done to assess the degree to which EMG variables could account for variance in the AIH-induced change in strength. More precisely, we analyzed whether relative changes in active region EMG amplitude and EMG area were able to predict changes in strength. We included the relative change in active region EMG amplitude and active region area, and their interactions as fixed effects. As random effects, we included random intercepts for each subject and contraction direction (i.e., flexion and extension). Variance accounted for by the model is reported as conditional R^2^GLMM values, whereas variance accounted for by only the fixed effects is reported as marginal R^2^GLMM values ^20–22^.

## Results

### AIH-induced effects on maximal elbow strength

Overall, maximal voluntary strength at the elbow increased substantially from PRE to 60mPOST in the AIH condition, but not in the SHAM. AIH effects on maximal voluntary elbow flexion and extension are shown in Fig. 2.

**Figure 2:**
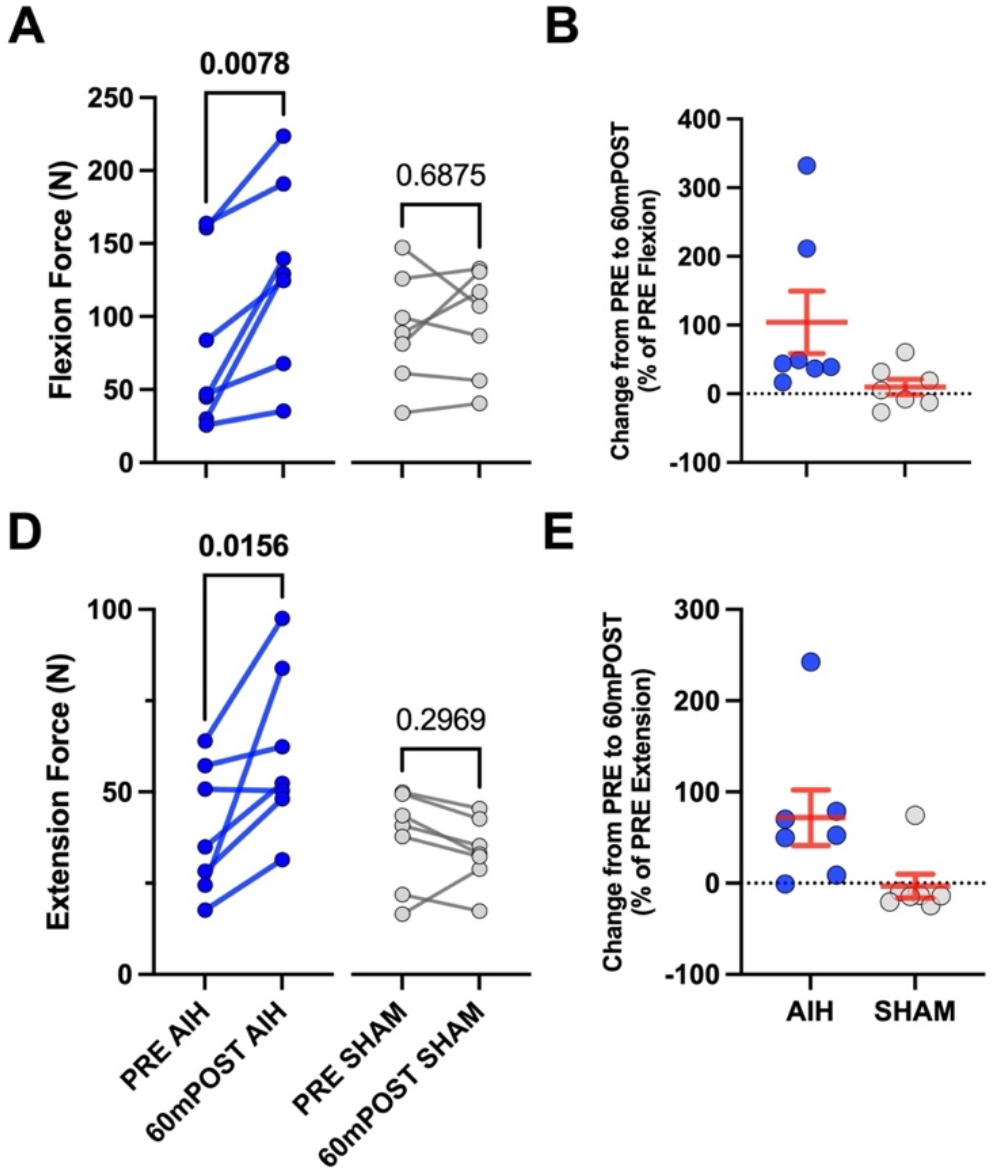
**Maximal elbow flexion (A) and extension (D) force of each participant during the AIH (blue) and SHAM (grey) conditions. P-values of the difference calculated with Wilcoxon matched-pairs signed rank tests are shown above each pre-post comparison. In B and E, the relative change in flexion and extension force, respectively, as a percentage of PRE is displayed for each participant with the blue (AIH) and grey (SHAM) points. The group mean ± standard error of the mean is shown in red**.

Elbow flexion strength did not differ between AIH and SHAM conditions at PRE (p = 0.8125) but increased by 50.7 ± 13.55 N (G = 0.58) from PRE (79.4 ± 22.55 N) to 60mPOST (130.2 ± 24.60 N) in the AIH condition (p = 0.0078). Furthermore, there was no change (p = 0.6875) from PRE (91.1 ± 14.37 N) to 60mPOST (95.9 ± 13.71 N) in the SHAM condition. The group mean of the relative increase in elbow flexion strength from PRE to 60mPOST AIH was 91.7 ± 33.5% of PRE and ranged from 16.4% up to 332.1%.

Elbow extension strength did not differ between AIH and SHAM conditions at PRE either (p = 0.08) but increased by 21.3 ± 7.57 N (G = 0.74) from PRE (39.6 ± 6.71 N) to 60mPOST (60.9 ± 8.57 N) in the AIH condition (p = 0.0156). Furthermore, there was no change (p = 0.2969) from PRE (50.4 ± 6.71 N) to 60mPOST (45.5 ± 4.73 N) in the SHAM condition. The group mean of the relative increase in elbow extension strength from PRE to 60mPOST AIH was 51.7 ± 21.9% of PRE and ranged from -0.9% up to 242.4%.

### AIH-induced effects on global EMG amplitude

Overall, muscle activity across the HDsEMG grid increased from PRE to 60mPOST in the AIH condition, but not SHAM.

During maximal elbow flexion, the global averaged root mean squared EMG amplitude of the biceps brachii increased by 141.1 ± 36.95 μV (G = 0.21) from PRE (650.0 ± 184.3 μV) to 60mPOST (791.0 ± 179.9 μV) in the AIH condition (p = 0.0313). Furthermore, there was no change (p = 0.6875) from PRE (639.7 ± 144.2 μV) to 60mPOST (671.6 ± 147.5 μV) in the SHAM condition. A representative EMG map is shown in Fig. 3 and AIH effects on global EMG amplitudes for each participant are shown in Fig. 4A.

**Figure 3:**
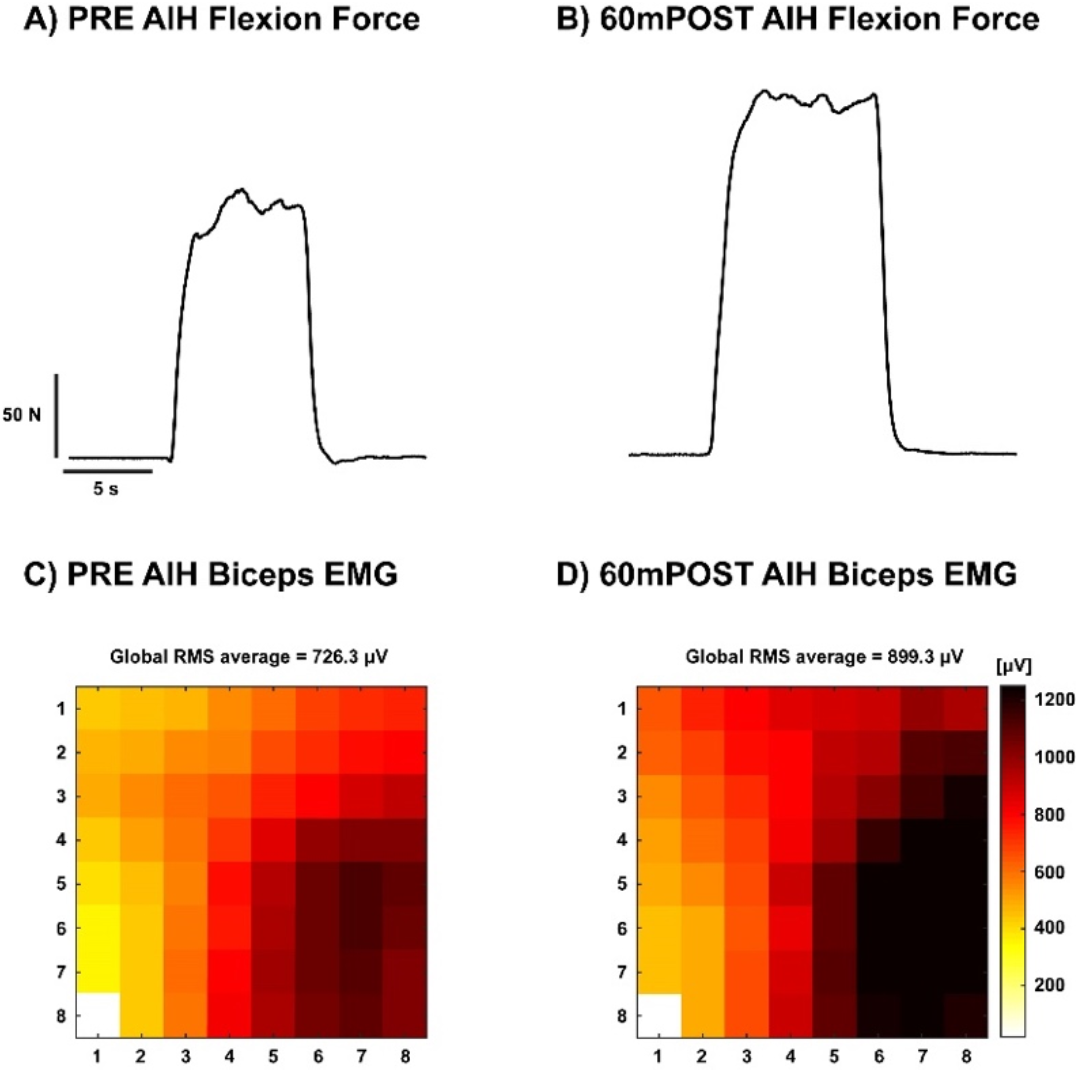
**A representative participant’s elbow flexion maximal voluntary force during PRE (A) and 60mPOST (B) in the AIH condition. Corresponding biceps brachii EMG maps during the contraction are displayed for PRE (C) and 60mPOST (D). Both EMG maps are displayed with RMS amplitudes (i.e. color intensity) on the same scale**.

**Figure 4:**
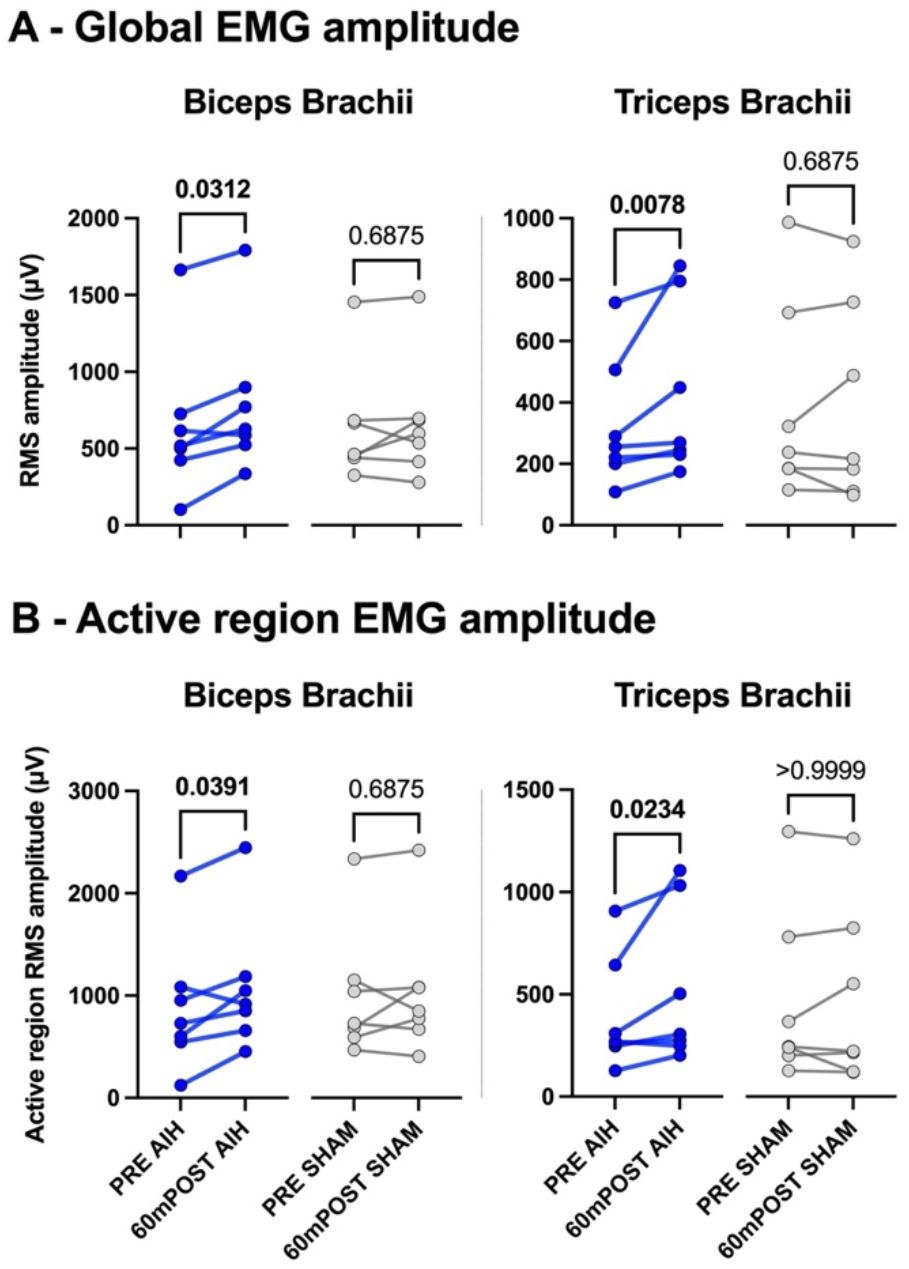
**Global (A) and active region (B) root mean square EMG amplitude in the biceps (left) and triceps (right) brachii during the AIH (blue) and SHAM (grey) conditions. *P-values of the difference calculated with Wilcoxon matched-pairs signed rank tests are shown in both A and B***.

Using watershed segmentation, we found that the activity of the active region of the HDsEMG grid was more intense after AIH, but not SHAM. During maximal elbow flexion, the EMG amplitude across the watershed segmented region of the biceps brachii increased by 193.7 ± 74.52 μV (G = 0.21) from PRE (886.6 ± 243.7 μV) to 60mPOST (1080 ± 245.3 μV) in the AIH condition (p = 0.0391). Furthermore, there was no change (p = 0.6875) from PRE (833.5 ± 200.4 μV) to 60mPOST (867.2 ± 205.6 μV) in the SHAM condition. AIH effects on active region EMG amplitudes for each participant are shown in Fig. 4B.

During maximal elbow extension, the global averaged root mean squared EMG amplitude of the triceps brachii increased by 100.5 ± 43.96 μV (G = 0.29) from PRE (329.4 ± 80.5 μV) to 60mPOST (429.9 ± 105.9 μV) in the AIH condition (p = 0.0078). Furthermore, there was no change (p = 0.6875) from PRE (389.3 ± 123.0 μV) to 60mPOST (392.6 ± 124.0 μV) in the SHAM condition. A representative EMG map is shown in Fig. 5 and AIH effects on EMG amplitudes averaged across the HDsEMG grid for each participant are shown in Fig. 4A.

**Figure 5:**
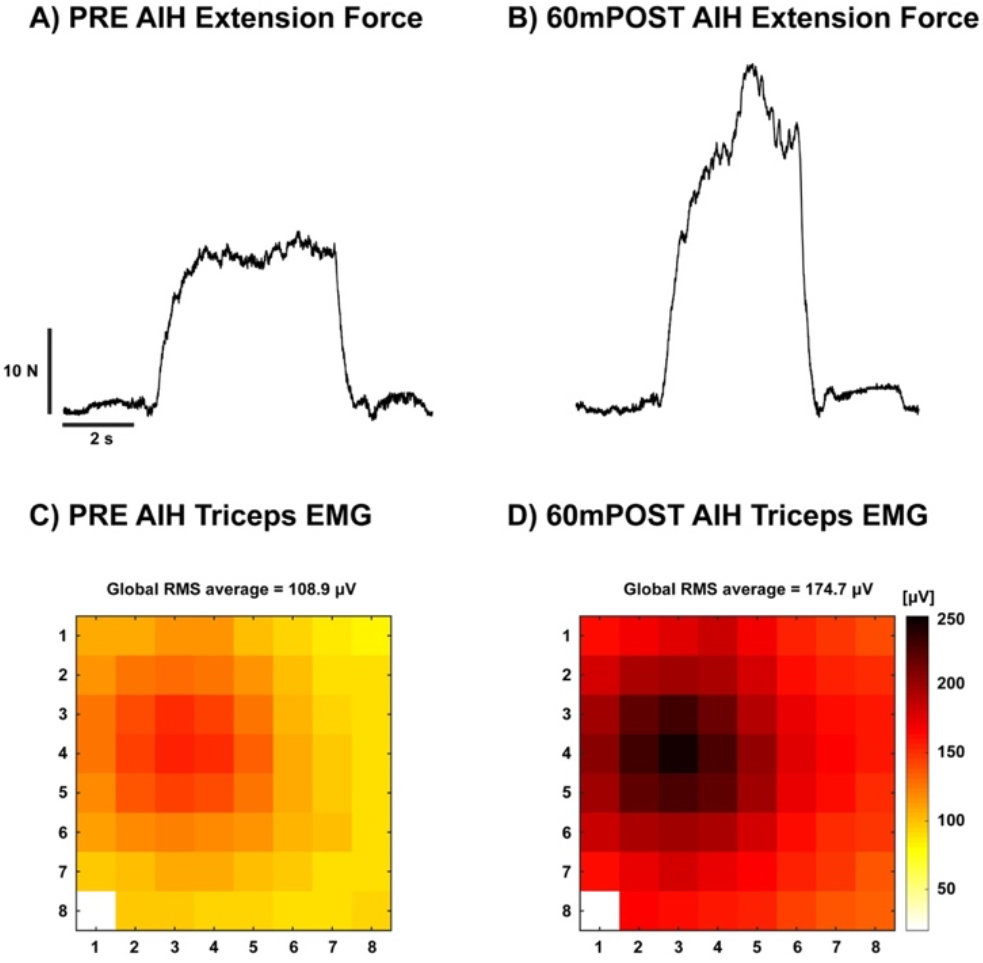
**A representative participant’s elbow extension force during PRE (A) and 60mPOST (B) in the AIH condition. Corresponding triceps brachii EMG maps during the contraction are displayed for PRE (C) and 60mPOST (D). Both EMG maps are displayed with RMS amplitudes (i.e. color intensity) on the same scale**.

Similarly, we found that during maximal elbow extension, the EMG amplitude across the watershed segmented region of the triceps brachii increased by 129.8 ± 61.65 μV (G = 0.28) from PRE (394.6 ± 104.4 μV) to 60mPOST (524.5 ± 145.3 μV) in the AIH condition (p = 0.0234). Furthermore, there was no change (p > 0.9999) from PRE (465 ± 160.6 μV) to 60mPOST (473.7 ± 163.8 μV) in the SHAM condition. AIH effects on active region EMG amplitudes for each participant are shown in Fig. 4B.

### AIH-induced effects on the spatial distribution of EMG activity

AIH effects on the area of the active region across the HDsEMG grid for each participant are shown in Fig. 6.

**Figure 6:**
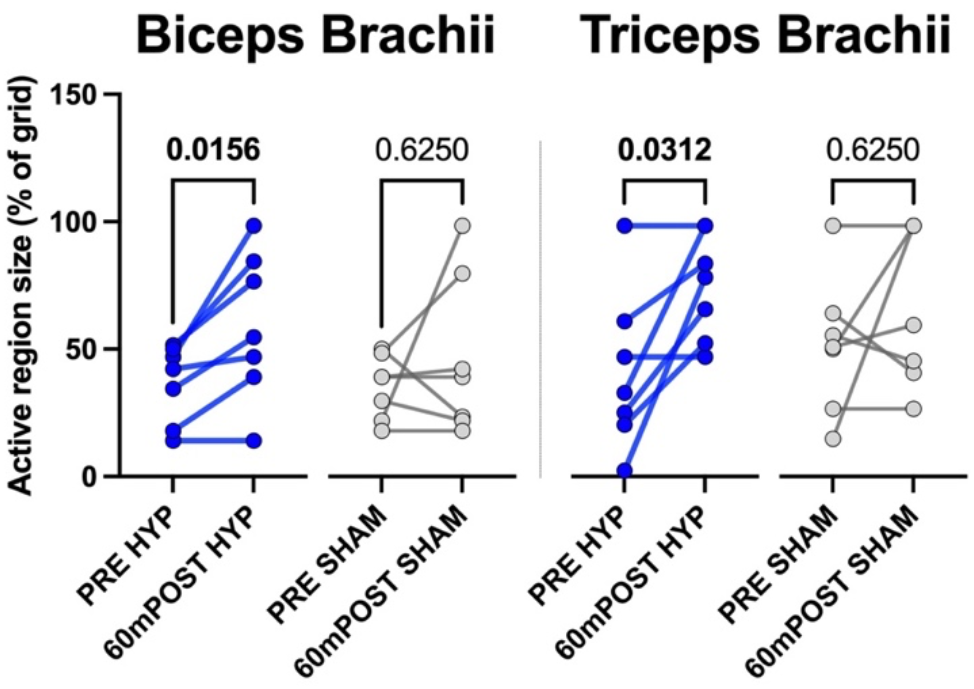
**The region of the biceps (left) and triceps (right) brachii EMG grid that exceeded 75% of maximum EMG amplitude of PRE during elbow extension of each participant during the AIH (blue) and SHAM (grey) conditions. P-values of the difference calculated with Wilcoxon matched-pairs signed rank tests are shown above each comparison**.

During maximal elbow flexion, the area of the biceps brachii grid with EMG amplitude exceeding 75% of PRE maximum increased by 22.4 ± 6.59% (G = 0.69) from PRE (36.7 ± 5.77%) to 60mPOST (59.2 ± 11.01%) in the AIH condition (p = 0.0156). Furthermore, there was no change (p = 0.6250) from PRE (35.2 ± 4.7%) to 60mPOST (46.1 ± 11.78%) in the SHAM condition.

During maximal elbow extension, the area of the triceps brachii grid with EMG amplitude exceeding 75% of PRE maximum increased by 33.8 ± 11.16% (G = 0.9) from PRE (40.9 ± 11.93%) to 60mPOST (74.8 ± 55.62%) in the AIH condition (p = 0.0313). Furthermore, there was no change (p = 0.6250) from PRE (51.5 ± 10.17%) to 60mPOST (66.7 ± 11.78%) in the SHAM condition.

### Relationships between AIH-induced effects on EMG activity and strength

We also determined whether any relationship existed between the relative change in EMG amplitude and area, and the AIH-induced change in strength. The generalized linear mixed model accounted for 84.6% of the observed variance in the AIH-induced change in strength, with the fixed effects accounting for 81.7% of the variance. Both the change in active region EMG amplitude (χ2 [2] = 23.2, P < 0.0001) and area (χ2 [2] = 9.847, P = 0.007275) were significant predictors of strength, but the interaction between them was not (χ2 [1] = 0.0994, P = 0.7526). Greater relative changes in strength were associated with larger increases in EMG amplitudes (1.33 ± 0.19, P < 0.0001) and area (0.05 ± 0.014, P = 0.00377). In summary, a positive relationship existed between changes in active region EMG amplitude and active region area, and changes in strength.

## Discussion

in addition to clarifying the underlying mechanisms, understanding the effects of AIH on elbow strength, in addition to the underlying mechanisms of this intervention, is an important step forward for improving neuroplasticity and functional recovery in people with SCI. Here we showed that AIH substantially enhanced maximum elbow flexion and extension strength in people with chronic incomplete cervical SCI. The increase in strength was accompanied by increases in the global average EMG amplitude, active region EMG amplitude, and the area of the active region across the EMG grid. Interestingly, the variance in AIH-induced increases in strength were predicted by changes in the amplitude and area of muscle activity. These findings suggest that AIH can improve the level of voluntary neural activation of the elbow flexors and extensors in individuals with incomplete SCI.

It was previously suggested that the relative magnitude of AIH-induced changes in strength (i.e., percent change) of the upper limbs (grip strength change of ∼10% from baseline) ^11^ is limited compared to that of the lower limbs (30-80% from baseline) ^8–10^. The current findings strike down that suggestion because we observed ∼90% and ∼50% increases in elbow flexion and extension forces, respectively. The relative magnitudes are in line with those observed in the plantarflexors (∼80%) ^8^, and may be linked to the functional roles of the muscle groups. Similar to the muscles of the lower limbs, proximal muscles of the upper limbs are well-suited for stabilizing and postural support of the limb compared to the distal muscles of the wrist and hand. These functionally distinct groups of muscles may have differences in motoneuronal properties (i.e., intrinsic motoneuron properties, relative distribution of synaptic inputs from various descending and spinal pathways, etc.) ^23^ that alter their susceptibility to AIH-induced changes in strength.

The mechanisms underlying AIH-induced neuroplasticity in respiratory motoneurons have been studied extensively in both human and non-human animals. In animal models, AIH triggers the release of serotonin, which leads to synthesis of brain-derived neurotrophic factor (BDNF) and activation of the high-affinity tropomyosin-related kinase B receptor ^5,12,15,16^. As a result, downstream signaling cascades enhance synaptic input onto spinal motoneurons ^4,13,14,17^.

Indeed, inputs from major descending tracts (i.e., the corticospinal tract) to the spinal motoneurons important for voluntary activation of muscle ^24^ are enhanced in both neurologically intact humans ^18^ and individuals with chronic incomplete cervical SCI ^25^. It is likely that the AIH-induced changes in strength observed in the current study could be attributed to similar mechanisms. Enhanced synaptic input would result in improved activation of motor units required for force generation.

The quantal elements for force generation are the motor units ^26^, which are comprised of the spinal motoneuron and all innervated muscle fibers. Modulation of muscle force can be achieved via two primary mechanisms: 1) recruitment of previously inactive motor units, or 2) increasing the discharge rate of active motor units. This study leaves us uncertain if the improved activation after AIH results from recruitment of new motor units or from increased firing rates of previously active motor units. Although both mechanisms promote increases in surface EMG amplitudes, the expansion of the active region as a result of AIH observed in the current study may point to recruitment of new motor units as a leading mechanism. On the contrary, increased EMG amplitudes of the active region may also suggest increased discharge rates of active motor units as the leading mechanism. Subsequent studies using advanced decomposition techniques ^27,28^ which will allow tracking of individual motor units from pre- to post-AIH may shed more light on the underlying mechanisms contributing to the observed increases in strength after AIH.

When we compare the maps of EMG activity emerging after AIH administration with those collected before AIH, there are clear and significant changes in the magnitude of EMG activity, but, although some changes in its spatial distribution were observed, shape changes in the EMG activity were less clear to determine. The resolution of our measurements is likely insufficient to determine whether the EMG map changes reflect a generalized increase in EMG activity at all active sites or whether there are also changes in the shape of the maps, suggesting that different motor unit populations may display a differential response to AIH. This distinction may require higher resolution HDsEMG measurements to support a definitive conclusion.

### Study Limitations and Future Implications

We acknowledge several study limitations and propose future directions based on our results. We recognize the impact of a small sample size of 7 subjects and the large variability in baseline strength among these subjects as a study limitation. Our data show that individuals showed an increase in elbow flexion strength from ∼16% to 300% suggesting that baseline strength likely affected the magnitude of plasticity after AIH, as has been previously reported ^9^.

A second limitation is that we did not test outcomes immediately after AIH until at least 60 minutes post-AIH. Future studies examining the time course of AIH-induced changes in muscle activation patterns may shed more light on different potential mechanisms mediating the improvement in strength after AIH.

We also did not test the effects of multiple sessions of AIH to determine whether there is an additive effect. Ultimately, our goal is to use AIH as an adjunct to rehabilitation to optimize functional tasks such as locomotor or upper limb function. The use of AIH to increase muscular force and activation shows great promise as a primer for improving rehabilitation outcomes. As suggested by previous studies ^1,29^, AIH should be combined with task-specific training to enhance functional outcomes. By doing so, AIH opens a window of opportunity for enhanced neural plasticity that can be exploited with task-specific training. Although the benefits of AIH in the current study are limited to ∼60 minutes post-AIH, it has previously been shown that some lasting effects on grip strength can be observed up to 24 hours after a single session ^11^. Future studies are required to determine whether repeated exposures to AIH can cause lasting improvements in strength and muscle activation that will help improve the quality of life in people with chronic SCI.

## Conclusions

These findings build on several reports from our lab, among others, demonstrating that AIH can improve motor function in people with chronic incomplete SCI. We show that strength and the spatial distribution of EMG activity of proximal muscles of the upper limb are enhanced following a single session of AIH. The magnitude of the effects of AIH on elbow flexion and extension strength are similar to those observed in the lower limb, but the change in the spatial distribution of EMG do not completely elucidate the underlying neuromuscular mechanisms. Further analysis of how AIH may affect individual motor unit behavior are needed to improve our understanding of AIH-induced changes in volitional strength.

## Data Availability

All data produced in the present study are available upon reasonable request to the authors

## Acknowledgements

The authors would like to extend thanks to Andres Cardona for his assistance with data collection, and to Alexander Barry for his help with ethical approval and clinical trial registration. The work was funded in part by a National Institute on Disability, Independent Living, and Rehabilitation Research (NIDILRR) training grant (GEPP), and a NIDILRR Midwest Regional SCI Model System grant (H133P110013; WZR and MSS).

